# MODIFIED AORTIC VALVE REIMPLANTATION IN PATIENTS WITH ACUTE AORTIC DISSECTION TYPE A

**DOI:** 10.1101/2024.02.07.24302455

**Authors:** Sergey Yuryevich Boldyrev, Valentina Nikolaevna Suslova, Mariya Vladimirovna Dontsova, Kirill Olegovich Barbukhatti, Vladimir Alekseevich Porkhanov

## Abstract

**Background:** The question of choosing surgical tactics in patients with acute aortic dissection type A unresolved to now. In patients with massive destruction of the aortic root the «gold standard» is the Bentall procedure. Aortic valve reimplantation is an attractive alternative, especially in patients with preserved aortic valve leaflets anatomy.

**Objective:** To compare the results of the use of valve sparing procedure and composite root replacement in patients with acute aortic dissection type A.

**Methods:** At the end, 62 patients were included in the study. Of those, 27 patients undervent aortic valve reimplantation and 35 had Bentall’s operation in the Kouchoukos modification.

**Results:** Preoperative demographic and clinical characteristics were such between the groups. Similar indicators of preoperative malperfusion were observed in both groups. The time of artificial circulation (p=0.125) and aortic clamping time (p=0.001) were longer in the reimplantation group, while the time of circulatory arrest was longer in the Bentall group (p=0.290). The average aortic clamping time was about 30 minutes longer in the first group. Hospital mortality in the reimplantation group was 8.3% and 22.9% in the Bentall group. Mortality from all causes in the long-term period was 6 people in the reimplantation group 25%, 10 people in the Bentall group 28.6%. The degree of aortic regurgitation did not increase in any case until the moment of extreme contact with the patient.

**Conclusions:** Aortic valve reimplantation can be performed with good immediate and long-term results in patients with preserved aortic valve leaflets with acute aortic dissection type A.

## INTRODUCTION

Acute type A aortic dissection of the ascending aorta is about to 80–90% of all acute aortic syndromes.^1^ Considering numerous literature sources, the ascending aorta dissection is found in approximately 2.5-5 per 100,000 people per year.^2,3^ Mortality risk is estimated as 1-2% per hour, and mortality without surgical treatment achieves 60%. Open surgical reconstruction of the ascending aorta is a life-saving operation, and it still remains a standard treatment for patients with acute type A dissection.^4^ Choosing surgical technique for this patients unresolved to now. Based on the above, it is pertinent to study the application of the technique of reimplantation of the native aortic valve, as well as the quality of life of patients in the immediate and distant postoperative periods.

## MATERIALS AND METHODS

The Ethics Committee of the State Budgetary Healthcare Institution “Research Institute - Regional Clinical Hospital No. 1 named after Professor S.V.Ochapovsky”, Health Care Ministry of the Krasnodar Region, approved this study, and it was done regarding the requirements of medical insurance.

## PATIENTS

We analyzed data from all patients with acute aortic dissection admitted to our center from 2003 to 2023. Performing required analyses we selected patients with type A acute aortic dissection who underwent urgent surgery. The patients who were undergone supracoronary ascending aorta represent were excluded. At the end, 62 patients were included in the study, they were identified and stratified by the risk of the initial condition severity. Of those, 27 patients underwent aortic valve reimplantation and 35 had Bentall-De Bono operation. The exclusion criteria for valve reimplantation were: pathology of the aortic valve flaps, fenestration presence, extremely severe hemodynamic condition of the patient.

## SURGICAL TECHNIQUE

In all cases, operative approaches were carried out through median sternotomy, the cannulation scheme was selected based on the dissection anatomy and according to the choice of the operating surgeon following the volume of the proposed aortic reconstruction. Preference for the arterial cannula placement was given mainly to the brachiocephalic trunk or the ascending aorta under ultrasound control. Hypothermic crystalloid cardioplegia was applied in all cases and antegrade cerebral perfusion was performed during circulatory arrest for brain protection. All patients underwent near-infrared spectroscopy (NIRS), measurements were carried out using the INVOS system (Somanetics). The hypothermia level was selected individually for each patient and ranged from 20°C to 26 °C.

Depending on a surgeon’s preferences, three variants of the reimplantation technique were applied: Seattle, David V\Miller, proper modification.

In our hospital, the reimplantation technique was started with the Seattle modification, which was relevant at that time, and only two patients had surgical repair. Selection of the vascular graft was performed by adding 3 mm to the fibrous ring, and the base of the vascular graft was trimmed in such a way to be positioned opposite the commissures, creating new sinuses.^5^

The David V/Miller technique was applied in five cases. Multiple (12-14) U-shaped sutures was placed on the fibrous ring. When selecting the size of the graft, 6-8 mm was added to the diameter of the fibrous ring, that left more space for the aortic valve leaflets. The vascular graft was narrowed in the proximal part by a polyester thread on the selected fibrous ring measuring tool. Further, the vascular graft plication was carried out on three sides with separate threads at the level of the sinotubular ridge in order to shape neosinuses.^6^

In our modification, the entire purposed intervention on the arc was primarily performed, and then artificial blood circulation was restarted and the work at the root was began.

Coronary arteries were exposed from abnormal or dissected tissue, leaving a 4-5-millimeter margin. During dissection assessment, the tissue structure and the presence of tears were evaluated. If dissection extended to the arterial ostia, the wall layers were juxtaposed with additional felt pads usage or plication.

Measurement of the aorta ring is carried out by inserting the original device into the left ventricle through the aortic ring, the size of which corresponds to one of the dimensions of the device working part.^7^ The vascular graft was selected as with the David V\Miller method according to the following formula: the size of the vascular graft = the size of the selected device + 8 mm. However, our method of narrowing the proximal part of the vascular graft include gardening it using two monofilaments horizontally streaches, retracting from the edge of the graft 1-2 mm, similar to a matters suture. This sutures are placed at a distance of 3 mm from each other in a checkered order. Finally, a margin of the proximal end of the vascular prosthesis with a width up to 5 mm was obtained. Then, only three U-shaped stitches were applied on the patches below the valve leaflets’ base, transmurally, from the inside out, with the needle and thread fixed to the prosthesis by stitching the base of the cuff between the horizontal stitches. After suturing the fibrous ring, the aortic valve was reimplanted and fixed inside the vascular prosthesis. Coronary buttons were sequentially implanted into the prosthesis.

Patients who did not match the inclusion criteria, had a traditional root replacement using a mechanical valve conduit according to the Bentall-De Bono method in the Kouchoukos modification.^8^ At the end of the operation, in most cases, biological glue was used to seal the anastomoses additionally.

## IMMEDIATE AND LONG-TERM RESULTS

Mortality caused by any cause and degree of the increasing regurgitation were issues of major interest. All patients with preserved native valves underwent annual routine examinations with echocardiography to assess the evolution of aortic regurgitation.

## STATISTICAL ANALYSIS

Data analysis was performed using statistical software SPSS 26 (IBM Corp., 2019, IBM SPSS Statistics for Windows, 26.0 version; IBM Corp., Armonk, New York, USA).

The statistical description of continuous quantitative variables is presented as the mean ± standard deviation, categorical variables were described by an absolute and relative frequency (abs. (% of n)). The David and Bentall-De Bono groups were compared regarding clinical signs: in the case with normally distributed continuous signs using the Student’s t-test for independent sample, and with continuous signs that do not match the normal law, using the Mann-Whitney U-test. Reviewing the data distribution for compliance with the normal law was assessed using the Kolmogorov-Smirnov test. Comparison of categorical (binary) feature frequencies was performed using the z-test for proportions

A p-value < 0,05 indicated statistically significant differences between the comparison groups. The Kaplan-Meier method was used to assess freedom from regurgitation over a 15-year period following surgery.

## RESULTS

Sixty-two patients were included in the present study, and 27 of those had aortic valve reimplantation according to one of the modifications. Table 1 presents the demographic and clinical characteristics of the patients. In both groups there were more males than females. The average age was 49 years. The body surface area of the patients in both groups demonstrated no significant difference (p 0,949). Besides, there was no significant difference regarding the presence of concomitant diseases in the groups. This study included patients with Marfan syndrome, in the reimplantation group there were 3 patients, and in the Bentall group - 9 patients. As well, there were patients with a bicuspid aortic valve, 1 patient was in the reimplantation group and 6 patients were in the Bentall group. A left ventricle ejection fraction was mostly preserved in both groups. In the second group the average diameter of the ascending aorta was larger than in the first group (p=0,036). Aortic arch aneurysm was observed in 4 cases, 1 - in the reimplantation group and 3 in the composite graft group.

**Table 1:** Preoperative variables according to group.

|  |  | Total<br>N=62 (%) | David<br>N=27 (%) | Bentall-DeBono<br>N=35 (%) | p Value |
| --- | --- | --- | --- | --- | --- |
| Sex | male | 48 (77,4) | 22 (81,5) | 26 (74,3) | 0,555<sup>a</sup> |
|  | female | 14 (22,6) | 5 (18,5) | 9 (25,7) |  |
| Age (y) |  | 48,98 ± 11,36 | 50,11 ± 10,2 | 48,11 ± 12,28 | 0,497<sup>b</sup> |
| S (m<sup>2</sup>) |  | 1,97 ± 0,24 | 1,98 ± 0,24 | 1,97 ± 0,24 | 0,949<sup>b</sup> |
| Concomitant | hypertension | 30 (48,4) | 10 (37,0) | 20 (57,1) | 0,119 |
|  | diabetes | 2 (3,2) | 2 (7,4) | 0 | 0,105 |
|  | hypertension & diabetes | 6 (9,7) | 1 (3,7) | 5 (14,3) | 0,165 |
| Marfan syndrome |  | 12 (19,4) | 3 (11,1) | 9 (25,7) | 0,152 |
| Bicuspid aortic valve |  | 7 (11,3) | 1 (3,7) | 6 (17,1) | 0,101 |
| Aortic annulus (mm) |  | 25,32 ± 2,33 | 25,00 ± 2,35 | 25,57 ± 2,32 | 0,343<sup>b</sup> |
| LV EF (%) |  | 51,74 ± 5,35 | 51,89 ± 5,32 | 51,63 ± 5,4 | 0,914<sup>c</sup> |
| Ascending aorta diameter (mm) |  | 60,61 ± 12,39 | 57,41 ± 11,61 | 63,09 ± 12,57 | 0,036<sup>c</sup> |
| Aortic arch aneurysm |  | 4 (6,5) | 1 (3,7) | 3 (8,6) | 0,440 |
| DeBakey type | I | 42 (67,7) | 18 (66,7) | 24 (68,6) | 1,0<sup>a</sup> |
|  | II | 20 (32,3) | 9 (33,3) | 11 (31,4) |  |
| Symptomatic malperfusion | cerebral | 5 (8,1) | 3 (11,1) | 2 (5,7) | 0,442 |
|  | renal | 2 (3,2) | 1 (3,7) | 1 (2,9) | 0,861 |
|  | coronary | 3 (4,8) | 1 (3,7) | 2 (5,7) | 0,718 |
|  | lower extremities | 2 (3,2) | 1 (3,7) | 1 (2,9) | 0,861 |
|  | renal & lower extremities | 1 (1,6) | 1 (3,7) | 0 | 0,255 |
| X-ray malperfusion | cerebral | 4 (6,5) | 3 (11,1) | 1 (2,9) | 0,197 |
|  | renal | 3 (4,8) | 2 (7,4) | 1 (2,9) | 0,418 |
|  | coronary | 4 (6,5) | 0 | 4 (11,4) | 0,072 |
|  | lower extremities | 2 (3,2) | 2 (7,4) | 0 | 0,105 |
|  | cerebral & renal | 4 (6,5) | 1 (3,7) | 3 (8,6) | 0,440 |
|  | cerebral & lower extremities | 1 (1,6) | 0 | 1 (2,9) | 0,376 |
|  | coronary & renal | 1 (1,6) | 1 (3,7) | 0 | 0,255 |
|  | cerebral & renal<br>& lower<br>extremities | 6 (9,7) | 4 (14,8) | 2 (5,7) | 0,233 |

<sup>a</sup> the exact Fisher criterion was used; <sup>b</sup> the Student's t-test was used for 2 independent samples; <sup>c</sup> the Mann-Whitney U-test was used; in other cases, the z-test was used to compare proportions; LVEF- left ventricular ejection fraction.

Totally, there were 42 patients with type 1 dissection, and in 20 cases there was type 2 aortic dissection. There was no significant difference in clinical and radiological manifestations of malperfusion.

Time of artificial circulation and aortic clamping was longer in the reimplantation group, while circulatory arrest time was longer in the Bentall group (Table 2). The mean time of the aorta clamping was about 30 minutes longer in the first group. All patients in the second group underwent implantation of a double-leaflets mechanical valve graft. In the reimplantation group coronary artery bypass grafting was performed in 3 cases with the extended dissection to the coronary ostia. Frequency of the complete aortic arch replacement was higher in the Bentall group. In the first group, repeated clamping was not required for further valve plasty.

**Table 2:**
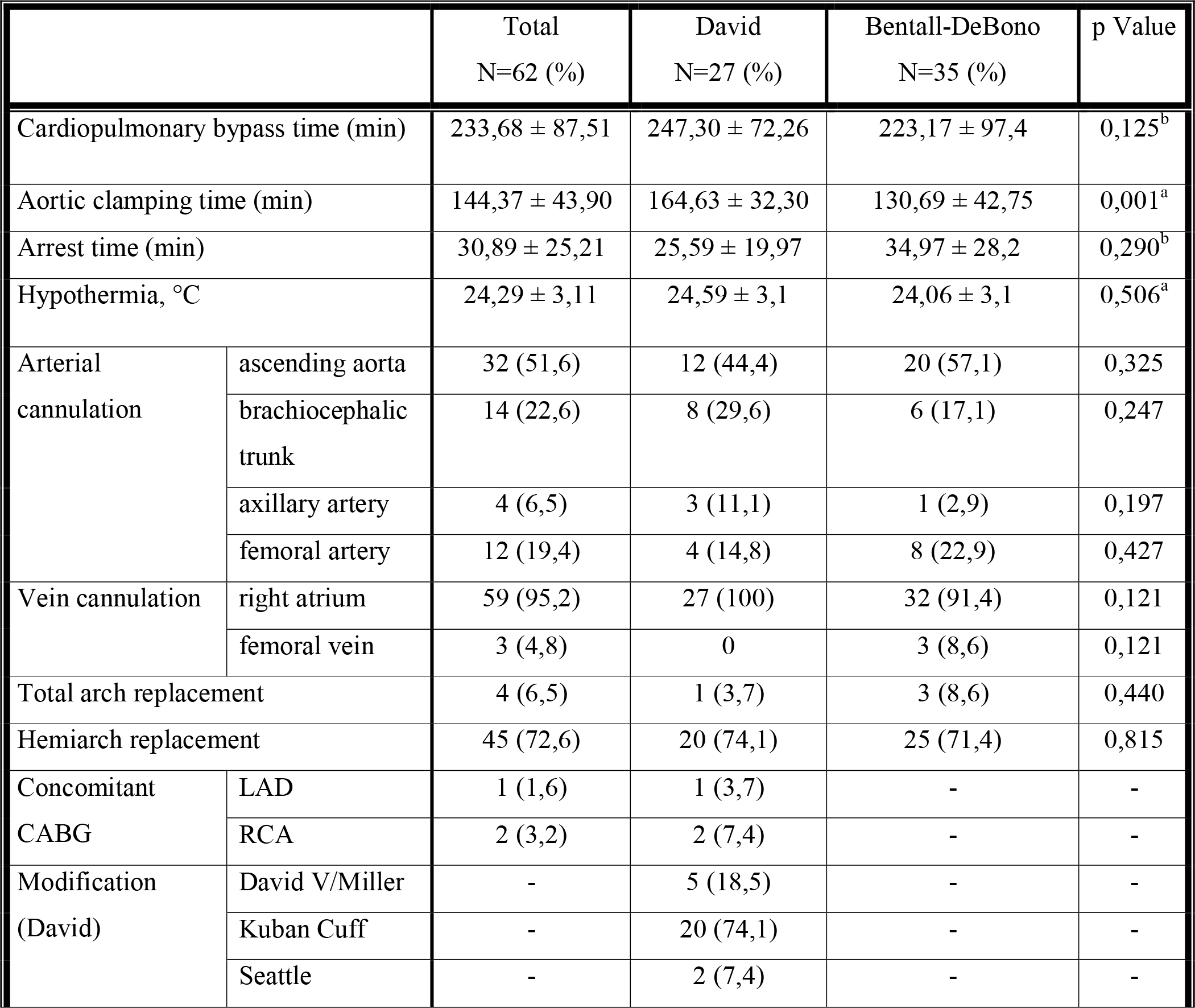
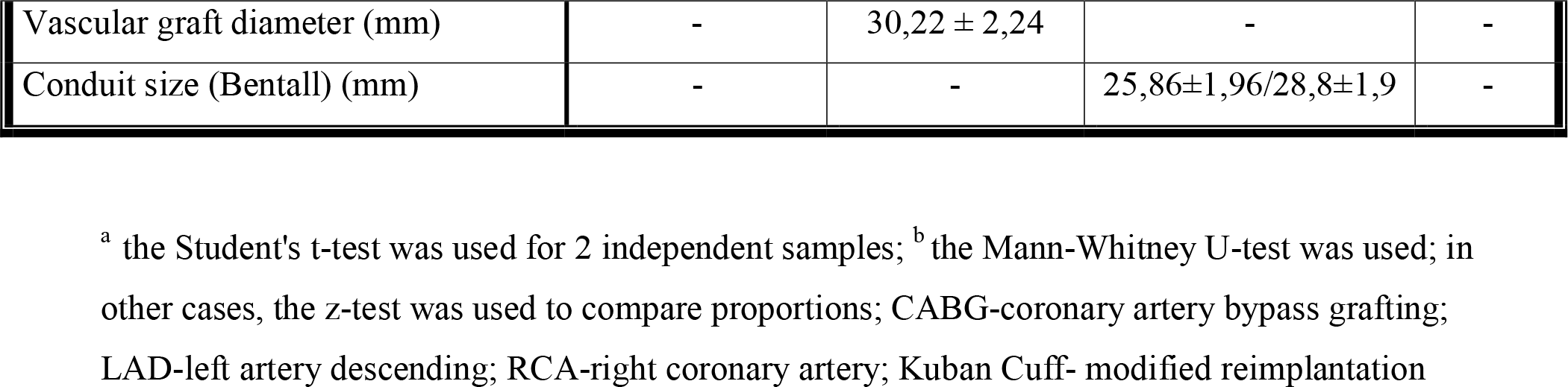
Operative variables according to group.

In the early postoperative period in the second group decreased cardiac output, requiring increased time for cardiotonic support, respiratory failure event requiring prolongation of artificial ventilation, cerebrovascular disorders, as well as re-sternotomy for further debridement on the first postoperative day were more common (Table 3).

**Table 3:** Postoperative outcomes according to group.

|  | Total<br>N (%) | David<br>N=27 (%) | Bentall-DeBono<br>N=35 (%) | p Value |
| --- | --- | --- | --- | --- |
| Low cardiac output | 11 (17,7) | 3 (11,1) | 8 (22,9) | 0,232<sup>a</sup> |
| Respiratory failure | 10 (16,1) | 4 (14,8) | 6 (17,1) | 0,809<sup>a</sup> |
| Cerebrovascular accident | 3 (4,8) | 1 (3,7) | 2 (5,7) | 0,718<sup>a</sup> |
| Reoperation for bleeding | 7 (11,3) | 2 (7,4) | 5 (14,3) | 0,399<sup>a</sup> |
| In-hospital mortality | 10 (16,1) | 2 (7,4) | 8 (22,9) | 0,103<sup>a</sup> |
| Total mortality | 16 (25,8) | 6 (22,2) | 10 (28,6) | 0,571<sup>a</sup> |

<sup>a</sup> the z-test was used to compare proportions

Hospital mortality rate was lower in the reimplantation group, with only 2 deaths comparing to 8 individuals in the Bentall group. One patient in the reimplantation group, diffuse tissue bleeding was observed as a result of developed consumption coagulopathy and subsequent myocardial infarction and the second patient required inotropic support in the postoperative period due to severe heart failure, which led to a fatal outcome.

Further follow-up of the patients showed that in the reimplantation group 2 more lethal cases occurred in the period up to 90 days after the operations. One patient died 78 days after the operation, pulmonary artery embolism was the cause of death. The second patient experienced acute heart failure 62 days after the operation. Mortality rate from all causes was lower in the reimplantation group comparing to the Bentall group (P =0,58; Fig. 1).

**Figure 1:**
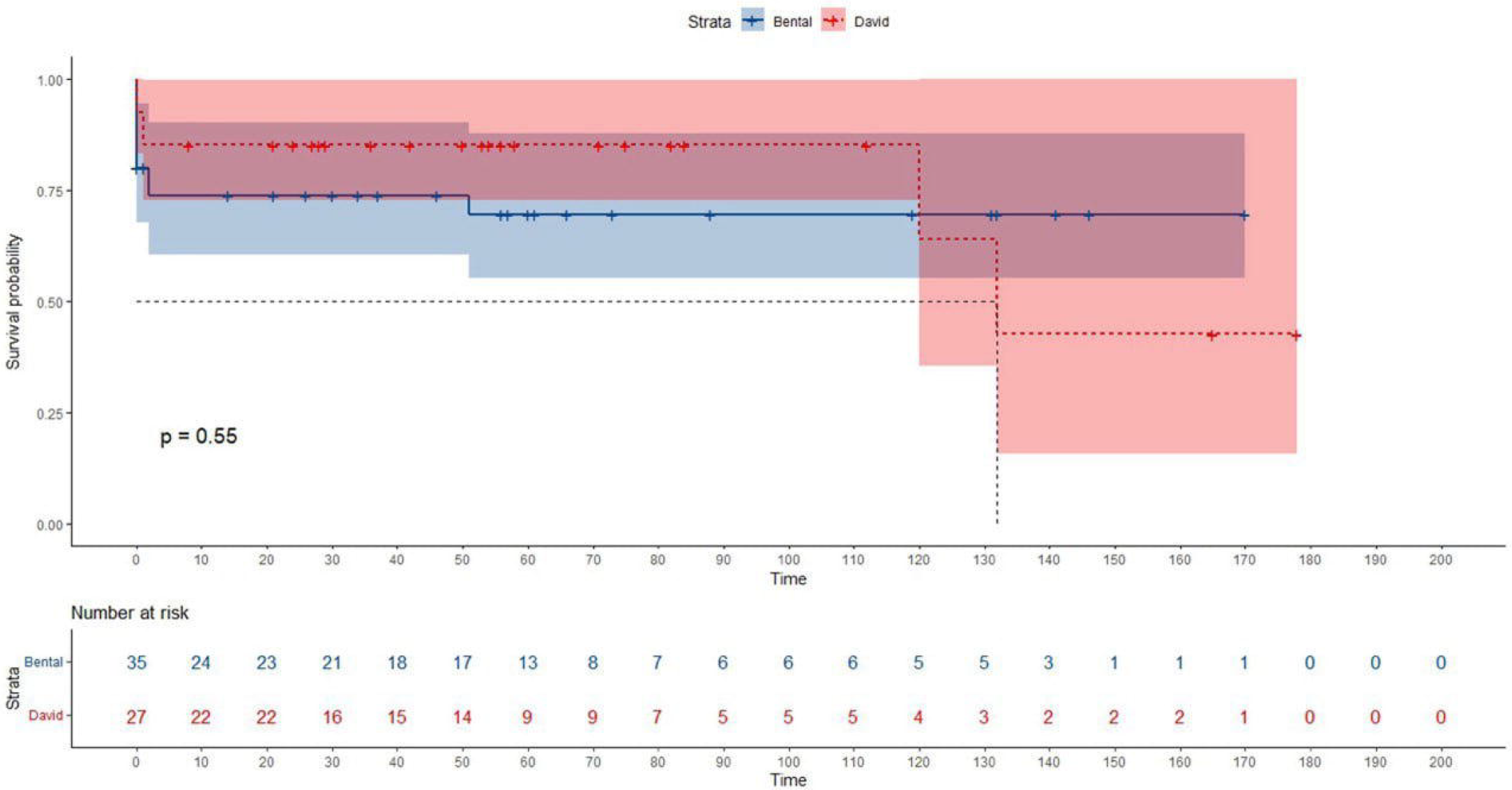
Analysis of the risk of death in the Bentall (blue) and David (red) group.

The degree of valve insufficiency did not increase in any case until the latest contact with the patient (Fig. 2).

**Figure 2:**
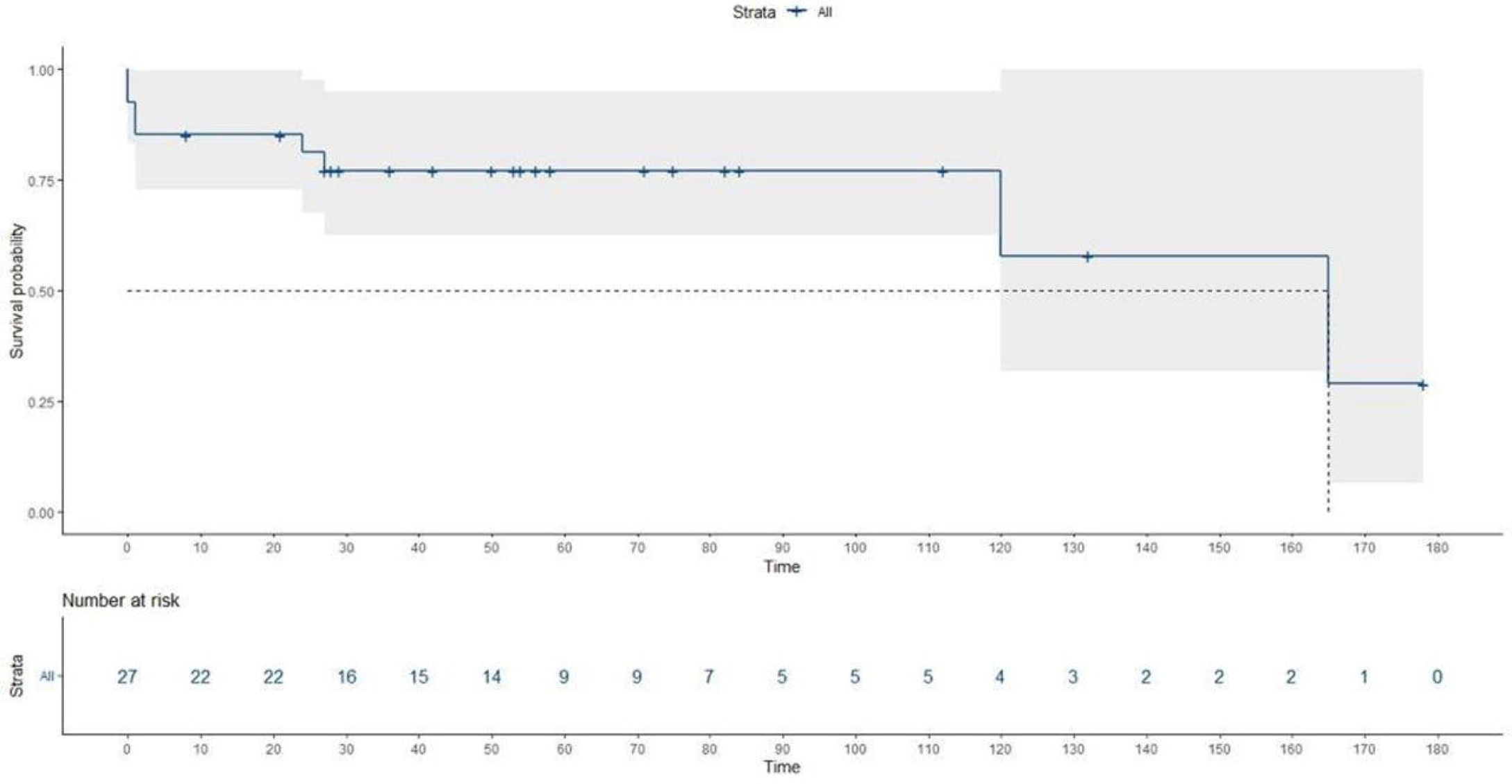
Degrees of aortic valve insufficiency in the postoperative period.

## DISCUSSION

According to the ACC/AHA guidelines for the diagnosis and treatment of aortic diseases and the AATS consensus on the surgical management of acute type A aortic dissection in most cases (evidence level IB) it is recommended to apply valve-sparing operations, except in cases with a dissection extending below the sinotubular junction.^4,9^ Supracoronary aortic replacement is recommended technique due to the reduced time of myocardial ischemia, artificial circulation and minimum dissection of altered tissues in patients in acute stage as it reduces risk of hemorrhagic complications. However, this technique reserves the risk for native root expansion in the future, and this event could require reoperation.^10,11^

Bentall De-Bono operation has proven its effectiveness for several decades, and now it remains the “gold standard” for type A aortic dissection due to its safety and feasibility (evidence level IB), however, the use of a mechanical valve condemns patients to lifelong anticoagulant therapy, potential hemorrhagic complications and an increased risk of thromboembolism.^4,12^

Considering that in most patients with dissection, aortic valve leaflets remain intact, techniques preserving the native valve have been proposed.^13,14^ In the Yakub remodeling technique, the lack of valve ring stabilization may lead to recurrent aortic insufficiency, and given the relatively long anastomotic line, it increases the risk of hemorrhagic complications, limiting the experience of performing this operation, especially in the context of tissue dissection.^15^

The revolutionary reimplantation technique initially described by David et al. in 1992, has advantages such as there is no need for long-term anticoagulant therapy with complete restoration of the aortic root and this making it a recommended treatment especially in young patients. When this technique is performed in expert centers, one could expect excellent outcomes without reoperations with a follow-up from 5 to 15 years.^16^

However, now, the use of aortic valve reimplantation procedures in cases with acute aortic dissection is limited.^17,18^ The International Register of IRAD reported 682 patients who underwent surgery in 18 centers, and aortic valve reimplantation was performed only in 5.8% patients.^19^ The German Registry of Acute Aortic Dissection Type A (GERAADA) presented data from 56 centers, showed that 8.2% were allocated for aortic valve reimplantation procedures.^20^ This fact is justified as the procedure of valve reimplantation is technically complicated, therefore, it is associated with prolonged operative time and requires extensive dissection of modified tissues, that in this pathology in the acute period can be a difficult task even for experienced surgeons.^21^ The combination of these factors dictates a specific tactical approach, particularly in patient selection for the reimplantation procedure in the context of acute aortic dissection.

In this study, we present a single-center experience in the surgical treatment for acute aortic dissection. We have been performing valve implantation according to the David method in our own modification as a priority manipulation.^22^ While using our technique, the number of sutures for graft fixation is reduced to three, and this fact quickens intervention at this stage. Also, for a better stabilization of the fibrous ring, two lines of sutures are placed when trimming the proximal part of the vascular graft. It should be mentioned that while using modified reimplantation, there were no cases with an increased degree of aortic regurgitation during the follow-up period for 15 years.

## CONCLUSIONS

The outcomes of the present study confirm the feasibility of surgical treatment for acute aortic dissection with preserved aortic valve leaflets anatomy using the reimplantation method in selected patients. Our results demonstrate excellent long-term survival and complete freedom from an increase in the degree of aortic regurgitation. We should emphasize that these types of operations for acute aortic dissection could be performed in expert centers by high-skilled surgeons.

## Data Availability

All data produced in the present work are contained in the manuscript

